# Association between Glucagon-Like Peptide 1 (GLP-1) Receptor Agonists Exposure and Intraocular Pressure Change

**DOI:** 10.1101/2024.05.06.24306943

**Authors:** Shahin Hallaj, William Halfpenny, Benton G. Chuter, Robert N. Weinreb, Sally L. Baxter, Qi N. Cui

## Abstract

**Objective:** This study aims to provide data on the effects of glucagon-like peptide 1 receptor (GLP-1R) agonists on intraocular pressure (IOP).

**Design:** Retrospective cohort study.

**Subjects, Participants, and/or Controls:** 1247 glaucoma surgery and treatment naïve eyes of 626 patients who were initiated on GLP-1R agonists compared to 1083 glaucoma surgery and treatment naïve eyes of 547 patients who were initiated on other oral antidiabetics.

**Methods, Intervention, or Testing:** The University of California Health Data Warehouse was queried for patients exposed to GLP-1R agonists or other oral antidiabetics. Index date was defined as the date of first exposure to the medication. Eyes with at least one pre-exposure and one post-exposure tonometry record within 365 days of the index date were included in the analysis. Clinical and laboratory data elements were extracted from the database. Eyes were censored from the analysis upon exposure to glaucoma hypotensive medication or glaucoma surgery. ΔIOP was analyzed using a paired t-test. Regression analysis was conducted using generalized estimating equations (GEE) accounting for inter-eye correlation. Sensitivity analyses were performed to assess the robustness of the findings.

**Main Outcome Measures:** Primary outcome measure was ΔIOP after exposure to the medication.

**Results:** The median age of all included subjects was 66.2 years [IQR=18.3]; 607 (51.7%) were female, and 667 (56.9%) were Caucasian. Median pre-exposure IOP, HbA1c, and BMI were 15.2 mmHg [IQR=3.8], 7.5 [IQR=2.4], and 29.8 [IQR=9.4], respectively. 776 individuals (66.1%) had diabetes, with the median number of active oral antidiabetics being 1.0 [IQR=1.0], and 441 (37.5%) being insulin users. Several pre-exposure characteristics significantly differed between the GLP-1R agonist and the control group. The mean ΔIOP was -0.4±2.8 mmHg (paired t-test p<0.001) and -0.2±3.3 mmHg (paired t-test p = 0.297) in the GLP-1R agonist and other antidiabetics groups, respectively. Pre-exposure IOP was the only independent predictor of ΔIOP in multivariable GEE. Sensitivity analyses yielded similar results.

**Conclusions:** Although GLP-1R agonists were significantly associated with a decrease in IOP in the paired analysis, they were not associated with ΔIOP in multivariable GEE. Moreover, the difference between the ΔIOP in the two groups was small. Future prospective studies following a standardized dose and delivery method may provide further insights.

## Introduction

The pathophysiology of glaucoma is complex, multifaceted, and not fully understood.^1–3^ It has been hypothesized that elevated intraocular pressure (IOP) and impaired retinal blood flow are initiating factors in glaucomatous optic nerve damage.^3^ The onset of retinal neurodegeneration in glaucoma may also be influenced by cumulative oxidative stress in conjunction with chronic mechanical and vascular stress.^4^ Oxidative stress is closely linked to cellular senescence, mitochondrial dysfunction, excitotoxicity, and neuroinflammation, all contributing factors to glaucoma progression.^5–7^ Similar age-related mechanisms could also contribute to trabecular meshwork dysfunction and elevate IOP in open-angle glaucoma.^8–12^

The incretin hormone glucagon-like peptide-1 (GLP-1) plays a pivotal role in glucose homeostasis.^13^ GLP-1 receptor (GLP-1R) agonists are long-acting GLP-1 mimetics first prescribed for controlling blood glucose levels in type 2 diabetes, a disease that has been proposed as a risk factor for glaucoma.^14^ GLP-1R activation has been linked to other beneficial effects, including weight loss, protection against nonfatal stroke and cardiovascular death.^15^ Multiple existing studies suggest a potential link between GLP-1R activation and protection against neuroinflammation in age-related neurodegenerative processes. Specifically, GLP-1R agonists initiate a signaling cascade that inhibits the production of proinflammatory cytokines and monocyte activation, contributing to the slowing of neurodegeneration.^16–18^ Preclinical studies identified similar mechanisms of neuronal death in glaucoma, whereby GLP-1R agonists ameliorate retinal inflammation and astrogliosis to rescue retinal ganglion cells. ^19–28^ Clinically, retrospective case-control studies have shown up to a 44% decrease in the hazard of a new glaucoma diagnosis in diabetic patients exposed to GLP-1R agonists compared to unexposed controls.^29,30^

Interestingly, a preclinical study investigating the effects of topically administered GLP-1R agonists demonstrated evidence of IOP reduction in a mouse model of ocular hypertension.^31^ In this study, we assessed the association between exposure to these medications and ΔIOP in humans by analyzing real-world data from the University of California Health Data Warehouse (UCHDW) as an initial step towards investigating whether existing preclinical findings of IOP modification can be translated into the clinic.

## Methods

### Data source

UCHDW was used for this study. This de-identified database uses the vendor-neutral and open-source Observational Medical Outcomes Partnership (OMOP) Common Data Model (CDM).^32^ It collates electronic health records (EHR) and medical claim information of 8.7 million patients seen across all University of California (UC) Health sites since 2012.^33^ This study was approved by the University of California San Diego Institutional Review Board, which also granted a waiver of informed consent.

### Glucagon-Like Peptide-1 Receptor Agonist Group

Eyes with at least one IOP record in patients who were initiated on a new GLP-1R agonist (i.e., exenatide, albiglutide, dulaglutide, lixisenatide, semaglutide, or liraglutide) were queried from the UCHDW (Figure 1). The index date was set to the date of the first GLP-1R agonist exposure, as recorded in the “drug_exposure_start_date” field of the OMOP “drug_exposure” table.^32^ Patients were required to be 18 years or older, have been exposed to a GLP-1R agonist, and have undergone at least one tonometry measurement both pre- and post-exposure within 12 months from the index date. Conversely, individuals under 18 years of age, without tonometry records within 12 months from the index date, who underwent IOP lowering procedures or surgeries before the index date, or who have a history of exposure to glaucoma hypotensive medications before the index date were excluded from the study.

**Figure 1.**
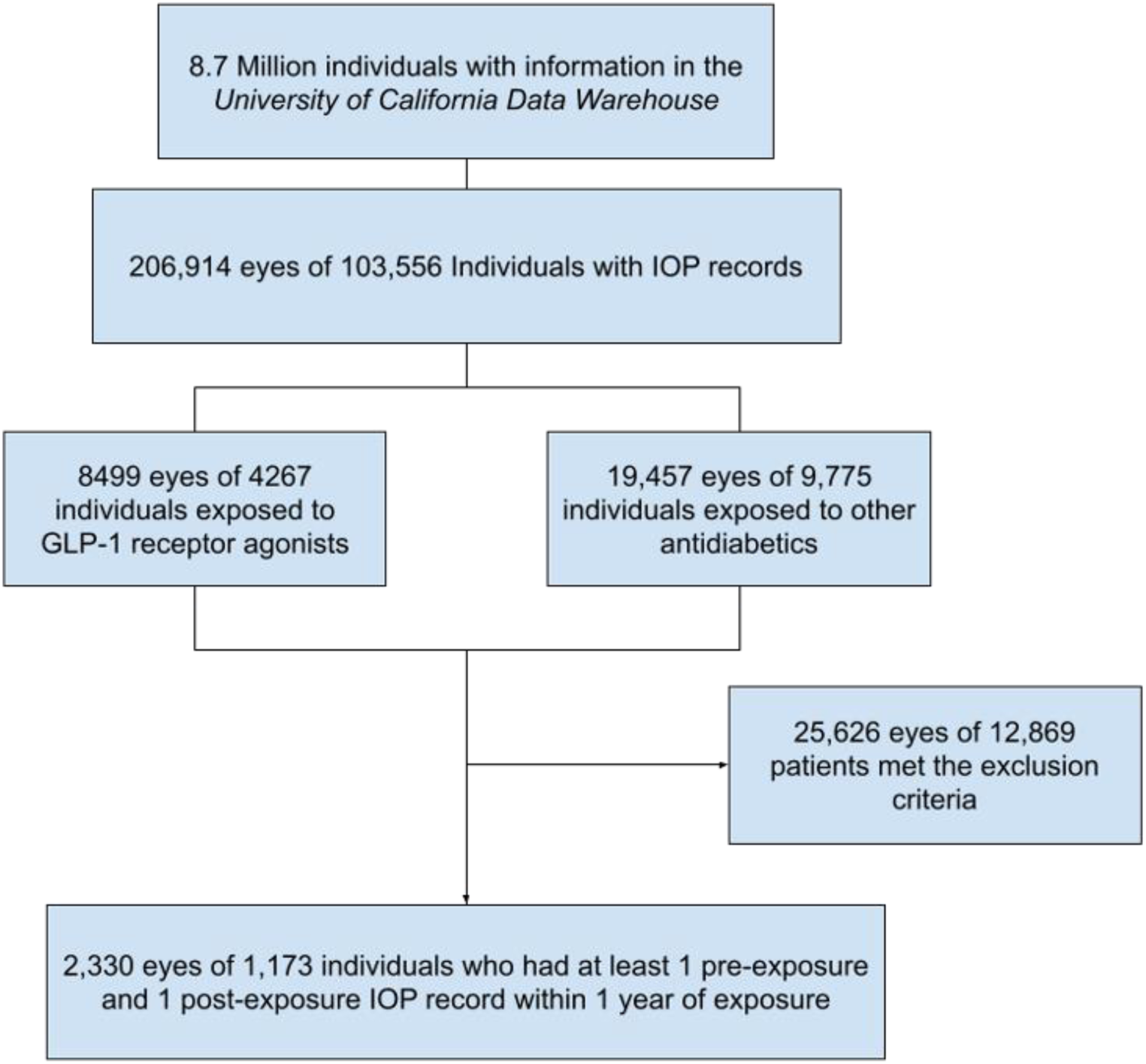
Flowchart depicting study cohort selection process. IOP: intraocular pressure, GLP-1R: glucagon-like peptide 1 receptor.

### Other Antidiabetics Group

The other antidiabetics group was comprised of patients not exposed to GLP-1R agonists, who initiated a new oral antidiabetic in one of the following subclasses: dipeptidyl peptidase 4 (DPP-4) inhibitors, sodium-glucose co-transporter 2 (SGLT2) inhibitors, sulfonamides, sulfonylureas, or thiazolidinediones. Eyes in this group were also required to meet all inclusion and exclusion criteria outlined above. This was chosen as the control group to allow consistency in methodology, e.g. analysis of IOP measurements around an index date linked to medication exposure. Eyes exposed to both GLP-1R agonists and other antidiabetics were included only in the GLP-1R agonist group. Only the first exposure to the medications was included in the analysis. Eyes were censored from the analysis if they were initiated on a hypotensive glaucoma medication or received an IOP lowering procedure or surgery. After applying inclusion and exclusion criteria, the subset of data available for this study included all IOP records in the database from November 4, 2017, to November 4, 2023. See Supplemental Table 1 for codes used in this study.

### Outcomes and covariates

The primary outcome measure was the difference between pre-exposure and post-exposure IOP (ΔIOP). Pre-exposure IOP was defined as the mean of the three most recent readings within 365 days before exposure. Post-exposure IOP was the mean of all readings within 365 days after exposure. The secondary outcome measure was the difference between pre- and post-exposure IOP variance. Demographic covariates assessed at the time of the index date were age, race, and gender. Ocular health factors included were a history of ocular hypertension or glaucoma suspect, a history of glaucoma, the stage of diabetic retinopathy (i.e., none, non-proliferative, proliferative, or unspecified), and history of intravitreal anti-vascular endothelial growth factor (VEGF) injections, as anti-VEGF injections have been associated with increased glaucoma risk.^29,34,35^ Systemic health state measures included histories of diabetes, hypertension, hyperlipidemia, and chronic kidney disease, body mass index (BMI), hemoglobin A1c (HbA1c) level, and the Diabetes Complications Severity Index (DCSI). The DCSI is a validated metric that is calculated using diagnosis codes from six categories of diabetic complications.^29^ Other covariates included insulin, statin, beta-blocker, and calcium channel blocker usage within 45 days from the index date.

### Statistical Analysis

The statistical analysis was conducted using the Python programming language (version 3.8.2, Python Software Foundation, Fredericksburg, Virginia, USA) and SQL queries on the UCHDW online Azure platform. When applicable, both eyes of each patient were included in the analysis. Descriptive analyses were conducted to evaluate the pre-exposure characteristics across the groups. The Shapiro-Wilk test was used to determine the normality of data distribution. Mann-Whitney U and Pearson’s Chi-squared tests were used to compare the pre-exposure characteristics between the two groups for continuous and categorical variables, respectively. To compare mean pre- vs. post-exposure IOP, a paired T-test was conducted, including the eyes of each group. A violin graph was used to demonstrate the changes in IOP distribution in each group. Linear regression models were fitted and graphed on a scatter plot to assess the correlation between the ΔIOP and pre-exposure IOP. Further subgroup analysis in eyes with glaucoma and glaucoma suspects was conducted using a paired t-test. Similarly, eyes were grouped based on their pre-exposure IOP using the median as the cutoff, and subgroup analysis was conducted using GLP-1R agonist exposure and pre-exposure IOP as predictors of ΔIOP in Generalized Estimating Equations (GEE) analysis. Adjustment for pre-exposure IOP was performed to control for regression to the mean. Univariable regression analysis was performed using GEEs to evaluate the associations between covariates and the outcome measure. GEE is an accepted, widely used modeling approach in ophthalmology literature, which allows accounting for inter-eye correlations when including both eyes of the patients.^36,37^ Variables with p<0.1 in the univariable GEE analysis were included in the multivariable GEE analysis. Sensitivity analyses were conducted to assess the robustness of the findings. P values <0.05 were considered statistically significant.

## Results

### Study Population

31584 tonometry records of 2330 eyes from 1173 individuals were included in the analysis. The overall cohort consisted of 1247 eyes of 626 individuals in the GLP-1R agonist group, with 1083 eyes of 547 patients in the other antidiabetics group.

### Patient-Level Characteristics

The median age of the included individuals was 66.2 years [IQR=18.3], with the greatest proportion being female (n=607 individuals, 51.7%) and Caucasian (n=667 individuals, 56.9%). Median pre-exposure IOP, HbA1c, and BMI were 15.2 mmHg [IQR=3.8], 7.5 [IQR=2.4], and 29.8 [IQR=9.4], respectively. 776 individuals (66.1%) had diabetes, with the median number of active oral antidiabetics being 1.0 [IQR=1.0], and 441 (37.5%) being insulin users. Several pre-exposure characteristics significantly differed between the GLP-1R Agonist and the control group, including age, gender, and race (Table 1).

**Table 1.**
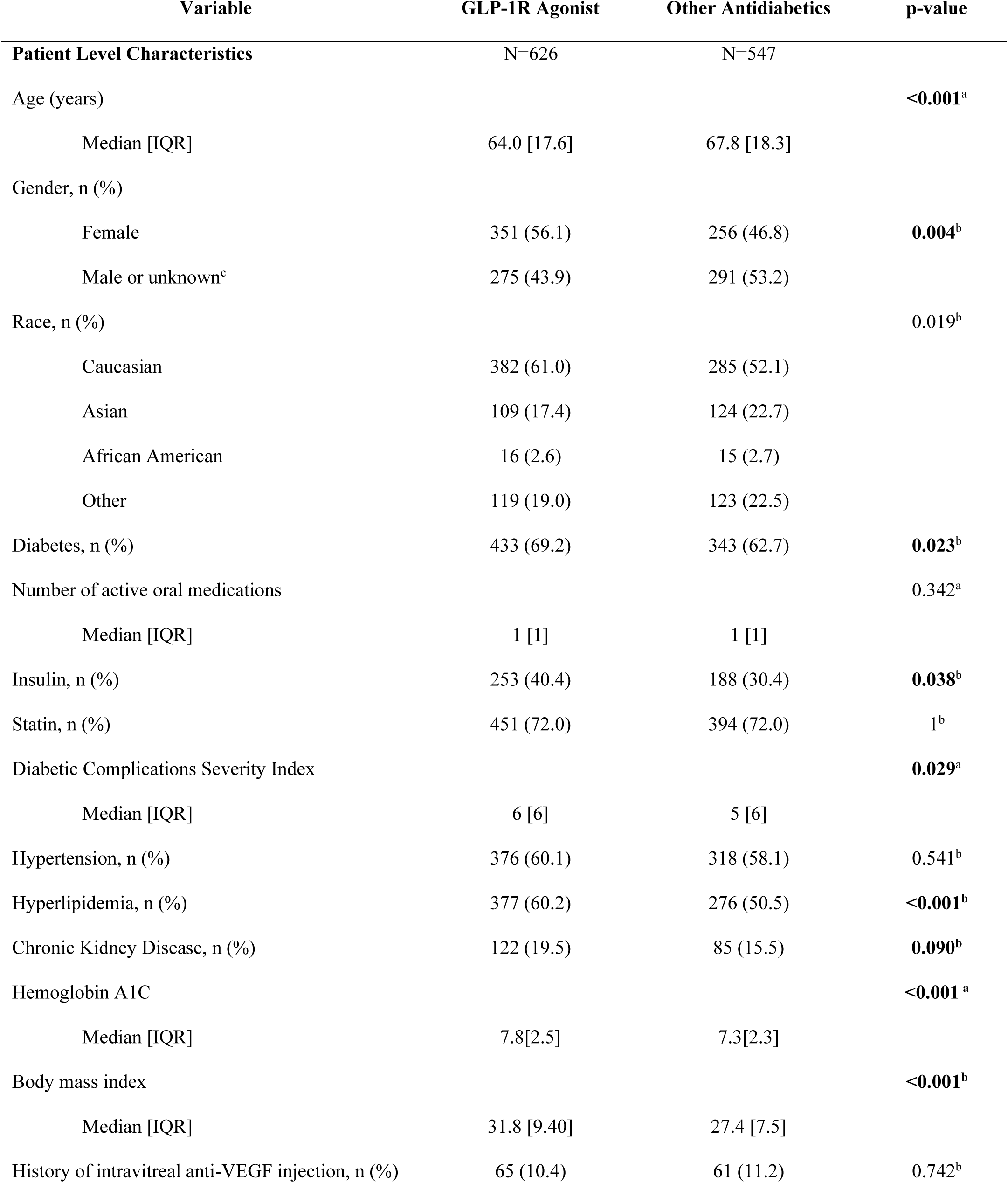
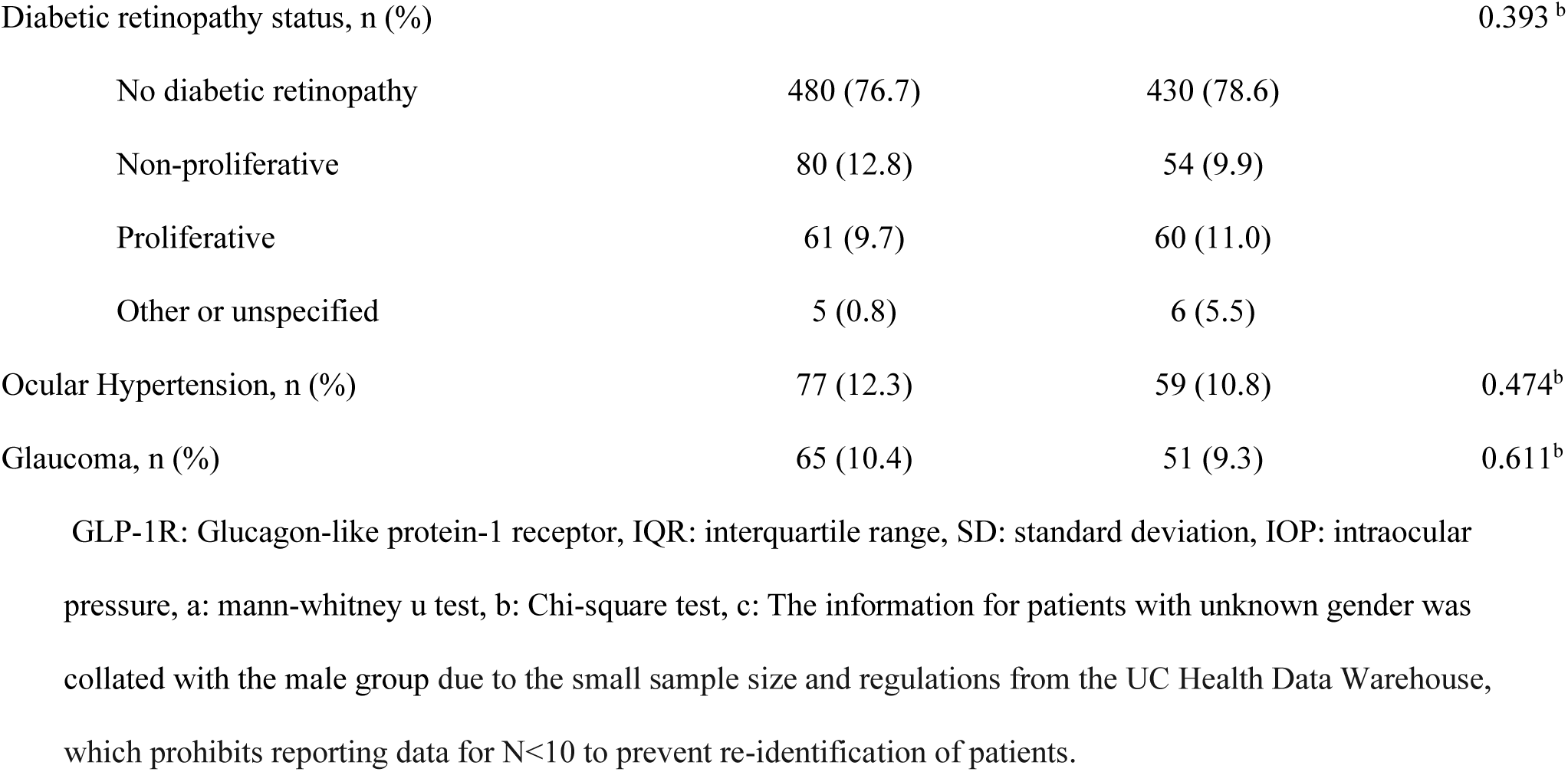
Pre-exposure characteristics of eyes in both groups.

### Eye-Level Characteristics

A diagnosis of glaucoma had been made in 231 eyes (19.7%), and 269 eyes (22.9%) were diagnosed as glaucoma suspects. While the mean number of tonometry records was similar between groups (p=0.080), eyes in the GLP-1R Agonist group had more pre-exposure tonometry records (median = 2.0 [IQR = 2.0] vs. 1.0 [IQR = 1.0], p=0.002) and fewer post-exposure records compared to those in the control group (median = 1.0 [IQR = 2.0] vs. 2.0 [IQR = 2.0], p=0.002). Table 1 provides further information about the differences between the groups.

### Observed Changes in Intraocular Pressure

The mean and median time periods between medication exposure and first IOP measurement were 106.4 and 71.0 days, respectively. The mean ΔIOP was -0.4±2.8 mmHg (paired t-test, p<0.001) and -0.2±3.3 mmHg (paired t-test, p = 0.055) in the GLP-1R agonist and other antidiabetics groups, respectively (Figure 2). In subgroup analysis, significant decreases in IOP post-exposure were observed in eyes treated with semaglutide, liraglutide, and SGLT2 inhibitors (Table 2).

**Figure 2.**
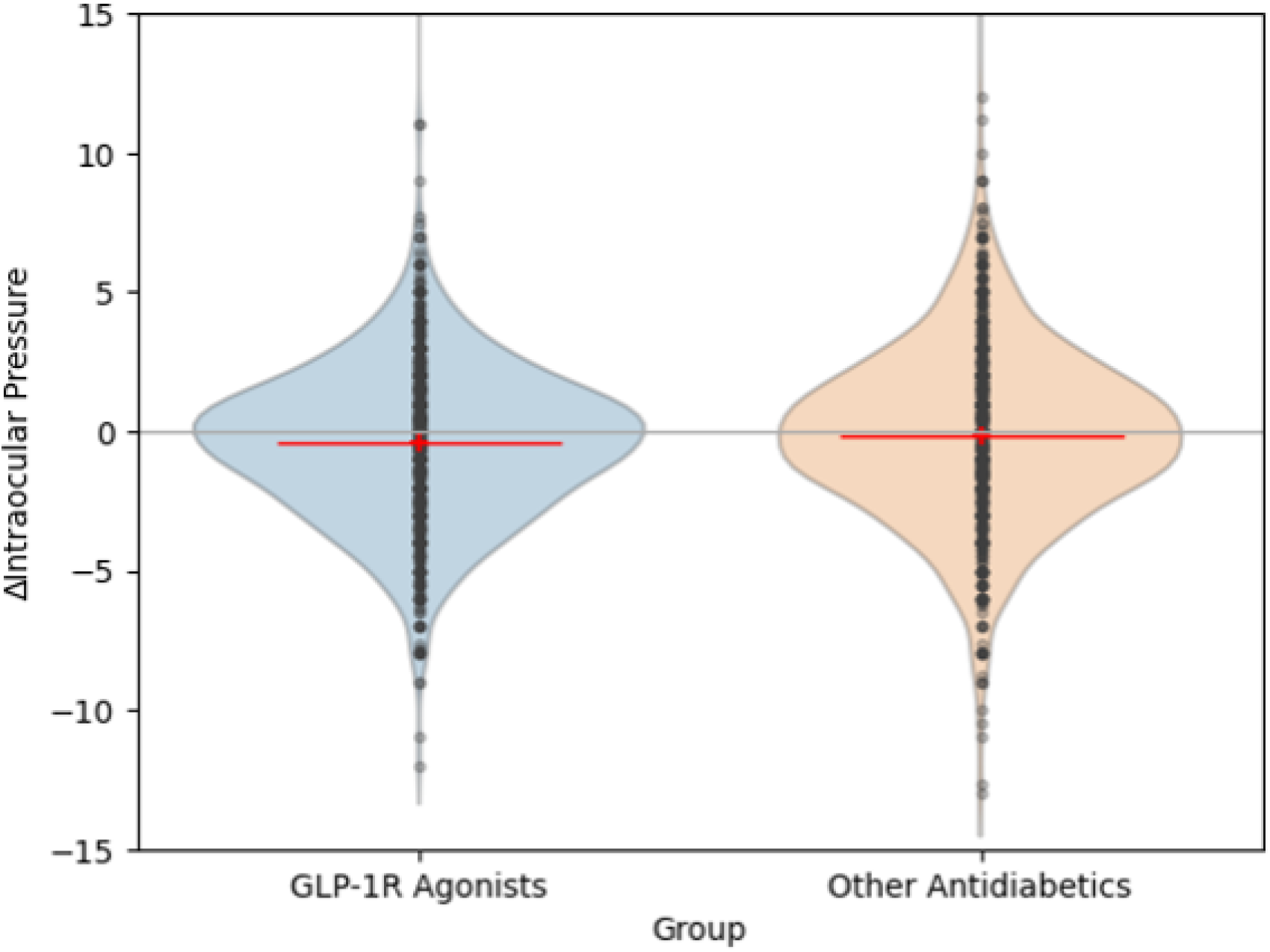
Violin plots comparing the distribution of Δ intraocular pressures (IOPs) between the groups. This change was only significant in the paired analysis of the individuals in the glucagon-like peptide 1 receptor (GLP-1R) agonists group.

**Table 2.** Changes in intraocular pressure pre- and post-exposure to multiple antidiabetic medication groups.

| Group | Number of eyes | Pre-exposure Median [IQR] | Post-exposure Median [IQR] | Mean Δ IOP | p-value<sup>a</sup> |
| --- | --- | --- | --- | --- | --- |
| All GLP1 receptor agonists | 1247 | 15.3 [4.0] | 15.0[4.0] | -0.4±2.8 | <0.001 |
| GLP1 receptor agonist subclasses<sup>b</sup> |  |  |  |  |  |
| semaglutide | 675 | 15.3 [4.0] | 15.0 [4.0] | -0.5 | <0.001 |
| Injectable | 517 | 15.3 [3.7] | 15[4] | -0.5 | <0.001 |
| Oral | 158 | 15.3[4.4] | 15[4] | -0.6 | 0.016 |
| dulaglutide | 375 | 15.3 [4.0] | 15.0 [3.9] | -0.3 | 0.06 |
| liraglutide | 137 | 16.0 [4.0] | 15.0 [3.4] | -0.6 | 0.006 |
| exenatide | 50 | 15.3 [3.5] | 15.5 [3.5] | 0 | 0.923 |
| All other antidiabetics | 1083 | 15.0[4.0] | 15.0 [4.0] | -0.2±3.3 | 0.055 |
| Other antidiabetics subclasses |  |  |  |  |  |
| Sodium-glucose co-transporter 2 (SGLT2) inhibitors | 485 | 15.0 [4.0] | 14.7 [5.0] | -0.4 | 0.005 |
| Dipeptidyl peptidase 4 (DPP-4) inhibitors | 273 | 15.0 [4.0] | 15.0 [4.0] | 0.1 | 0.756 |
| Sulfonylureas | 246 | 15.0 [4.0] | 15.0 [4.0] | -0.1 | 0.513 |
| Thiazolidinediones | 79 | 16.0 [4.5] | 15.0 [3.1] | -0.2 | 0.628 |

SGLT2: sodium-glucose co-transporter 2, DPP-4: dipeptidyl peptidase 4, SD: standard deviation, IQR: interquartile range, a: paired T test. b: An additional subclass, albiglutide, is not reported here due to the small sample size and regulations from the UC Health Data Warehouse, which prohibits reporting data for N<10 to prevent re- identification of patients.

ΔIOP was significantly correlated with pre-exposure IOP in both the GLP-1R agonist and control groups (Figure 3). Eyes with a pre-exposure IOP ≥ 15.0 mmHg had an average ΔIOP of -1.44 mmHg (p<0.001), whereas eyes with a pre-exposure IOP <15.0 mmHg had an average ΔIOP of +0.94 mmHg (p<0.001). The supplemental figure shows the regression of ΔIOP against the pre-exposure IOP. In the group with pre-exposure IOP above 15.0, the decrease in IOP trended towards being greater for the GLP-1R agonist group than for the other antidiabetics group (coeff= - 0.26, p=0.136) when adjusted for pre-exposure IOP. In the group with pre-exposure IOP below 15.0, the decrease in IOP trended towards being greater for the GLP-1R agonist group than for the other antidiabetics group (coeff= - 0.02, p=0.879) when adjusted for pre-exposure IOP. Subgroup analysis of ΔIOP based on glaucoma diagnosis is summarized in Table 3.’

**Table 3.**
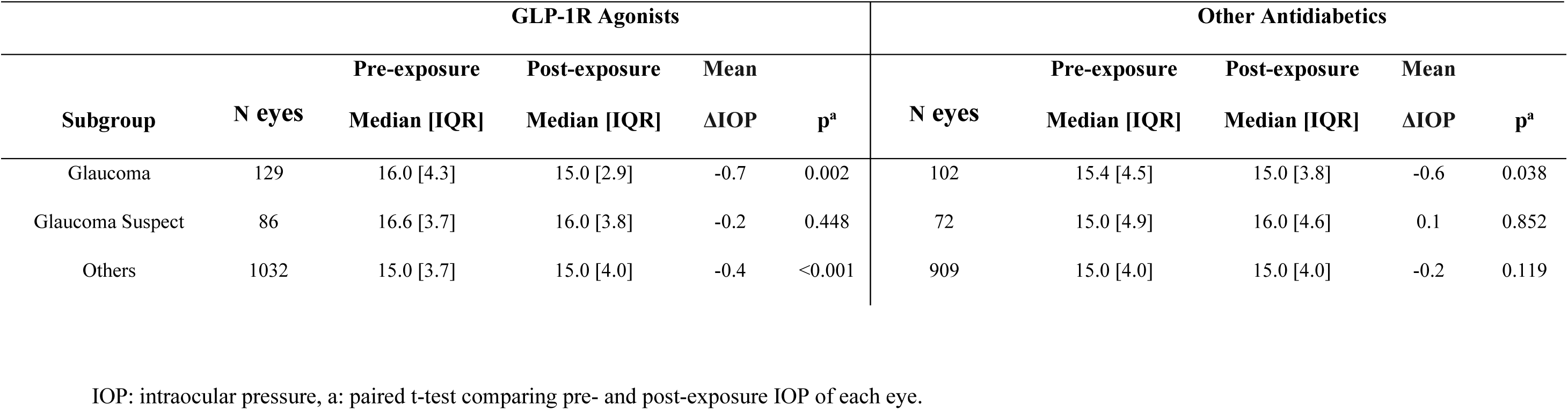
Comparison of pre- vs. post-exposure intraocular pressure (IOP) in glaucoma and glaucoma suspect eyes.

### Effect on Intraocular Pressure Variability

Filtering eyes with at least 3 pre-exposure and 3 post-exposure tonometry records resulted in a sample of 8738 tonometries performed on 327 eyes of 167 patients. Median pre-exposure IOP standard deviations in the GLP-1R agonist and control groups were 3.0 mmHg [IQR=6.0] and 3.0 mmHg [IQR=7.0], respectively. There were no significant differences in IOP standard deviation post-exposure, with GLP-1R agonist group having a post-exposure IOP standard deviation of 5.0 mmHg [IQR=5.5; p=0.197] and the control group with 5.1 mmHg [IQR=6.0; p=0.192]. Semaglutide (p=0.512), liraglutide (p=0.303), and SGLT2 inhibitors (p=0.237) were not significantly associated with changes in IOP variance post-exposure.

### Generalized Estimating Equations

Although the paired analyses demonstrated that GLP-1R agonist exposure was associated with a significant decrease in IOP, in a multivariable regression model with GEEs accounting for inter-eye correlation and other covariates, GLP-1R agonists were not independently associated with ΔIOP (Table 4). Pre-exposure IOP and hypertension were independent predictors of ΔIOP following exposure to antidiabetics. These results were robust to the sensitivity analyses of different medication subgroups, yielding similar results.

**Table 4.**
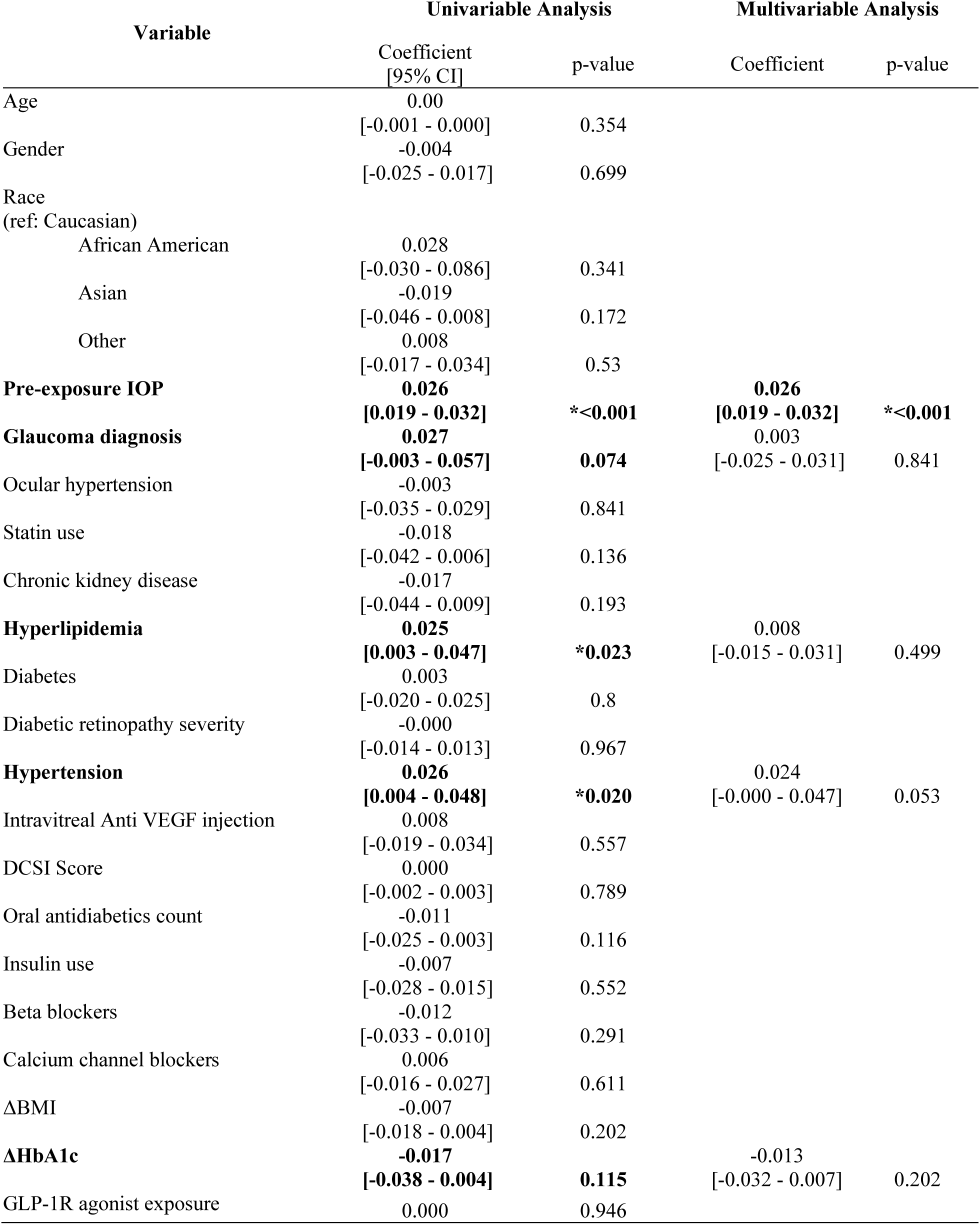
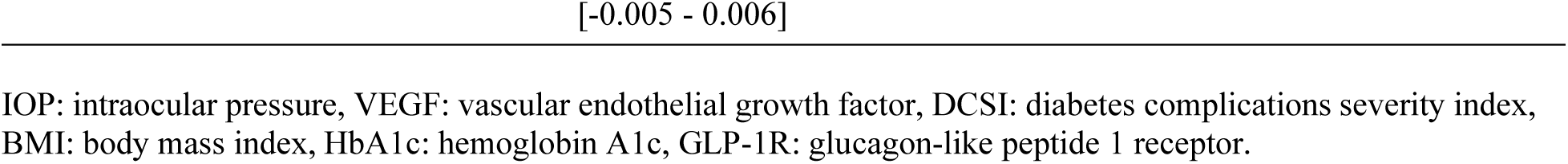
Generalized Estimating Equation Analysis of Δ Intraocular Pressure (mmHg) Accounting for Inter-eye Correlation.

## Discussion

There has been a growing interest in examining the effects of GLP-1R agonists for treating glaucoma. Neuroprotective effects of this class of medications have been shown in pre-clinical models of ocular diseases with a neurodegenerative component, including glaucoma.^38,39^ GLP-1R agonists have been shown to mitigate neuroinflammatory processes and offer protection against glaucomatous neuronal damage.^31^ Two large-scale retrospective case-control studies both reported a lower risk for a new glaucoma diagnosis in patients taking GLP- 1R agonists compared to other antidiabetic medications.^29,30^ Inspired by evidence of IOP lowering in a mouse model of hypertensive glaucoma exposed to GLP-1R agonists, this report is among the first to examine the effect of GLP-1R agonists on IOP. We reported the following main findings, to be discussed in turn: (1) a small, but statistically significant decrease in IOP following GLP-1R agonist initiation using paired t-test analysis, (2) an IOP decrease that is more prominent among eyes with pre-exposure IOPs ≥ 15 mmHg and among those with a history of glaucoma, and (3) GLP-1R agonist exposure was not an independent predictor of IOP in multivariable regression models when other covariates were included.

First, a significant decrease in IOP was noted in the GLP-1R agonist group following medication initiation, whereas a significant change was absent in those initiated on another antidiabetic agent. Among GLP-1R agonists, semaglutide and liraglutide, the most frequently initiated GLP-1R agonists in our cohort, were associated with a significant change in IOP. Interestingly, liraglutide and oral semaglutide were associated with the greatest decrease in IOP despite their comparatively smaller numbers, a finding at least in part consistent with previously reported preclinical data.^31^ Injectable and oral formulations of semaglutide demonstrated similar levels of ΔIOP. It is worth noting, however, that the median IOP decrease following agonist exposure was 0.4 mmHg, and may not be clinically relevant. This limited effect size may be attributable to variabilities in the route and dosage of administration not captured by queries of the database. Although several pre-exposure characteristics differed between the two groups, it should not affect the results of the paired t-test analyses, as the paired t-tests involve analyzing the distribution of changes in IOP for each individual before and after the exposure, rather than comparing population-based means. Another class of medication that was associated with ΔIOP in this study was SGLT2 inhibitors. Chieh Shao et al. found that SGLT2 inhibitors reduced incident glaucoma over GLP-1R agonists in a large multi-institutional cohort of type 2 diabetic patients of Asian descent.^40^ In comparison, while our study demonstrated that SGLT2 inhibitors slightly lowered IOP, ΔIOP was greater in the GLP-1R agonist group compared to the SGLT2 group.

Second, ΔIOP demonstrated a significant correlation with pre-exposure IOP, with the most notable change observed in eyes with a pre-exposure IOP of 15 mmHg or greater. Within this subgroup, exposure to GLP-1R agonists resulted in a mean ΔIOP of -1.44 mmHg, suggesting a potentially clinically significant IOP reduction in this subgroup (Table 3). This is consistent with preclinical data, which demonstrated IOP decrease in ocular hypertensive eyes but not in eyes without elevated IOP following GLP-1R agonist administration.^31^ Further, ΔIOP was more prominent in glaucomatous eyes. In this study, we examined all individuals exposed to GLP-1R agonists to leverage the largest sample size possible for our statistical analyses. However, based upon these findings that showed a modest reduction among “all-comers” but a more substantial (and potentially clinically significant) reduction among certain subgroups, future analyses in specific subgroups such as ocular hypertensive and glaucomatous eyes are merited.

Last, when compared to controls consisting of exposure to other antidiabetic medications, GLP-1R agonists were not independent predictors of IOP reduction in multivariable GEE models that adjusted for additional covariates. However, several considerations should be noted when interpreting these results. First, the paired analysis showed that *within* the GLP1-R agonist group, there was a statistically significant reduction in IOP after exposure, but the multivariable GEE demonstrates that this difference was not statistically significant when comparing *across* groups (i.e., comparing the GLP1-R agonist group to the controls exposed to other antidiabetic medications). One possibility is that in the multivariable model, other factors were stronger drivers of IOP reduction. For example, our multivariable GEE suggests that pre-exposure IOP may be strongly associated with ΔIOP following exposure to antidiabetics. We investigated different cohorts and accounted for different confounding variables in the sensitivity analyses, all of which confirmed the primary results in the general population. There may also have been additional confounding factors that were not accounted for in our models, such as variations in prescription patterns, patient preferences, medication tolerability, and other factors that are not easily discerned from EHR data and outside the scope of this study. Finally, similar to our previous comments addressing subgroup analyses by pre-exposure IOP and glaucoma status, it is possible that multivariable GEEs within specific groups (i.e., those diagnosed with glaucoma) may yield different results. A known confounding factor when dealing with longitudinal IOP data is regression to the mean^41^, a phenomenon that has been reported even in prospective landmark clinical trials such as the ocular hypertension treatment study (OHTS). ^42^ The observed trend in ΔIOP may have been in part due to regression to the mean. Aiming to account for this possibility, we ran a GEE analysis adjusting for pre-exposure IOP, showing that GLP-1R agonist exposure may be associated with a greater IOP decrease, a correlation that did not reach statistical significance but should be further explored in future studies.

These results broadly align with previous knowledge of GLP-1R agonists gained from preclinical studies.^31^ A clue hinting at a possible mechanism for GLP-1R agonist-associated IOP reduction lies in its recent evaluation as a medical option for managing idiopathic intracranial hypertension.^43^ Specifically, GLP-1R agonists have been shown to decrease cerebrospinal fluid production by inhibiting Na+/K+ ATPase activity within the choroidal plexus.^44^ Aqueous humor secretion is similarly dependent upon Na+/K+ ATPase activity, and if GLP-1R agonists directly inhibit Na+/K+ ATPase within the ciliary body, this is one pathway through which aqueous secretion and, by extension, IOP might be decreased. Although weight loss, a known and desirable effect of GLP-1R agonist use, could play a role in IOP determination,^45^ in our study, ΔBMI did not show any association with ΔIOP. However, with only 935 ΔBMI data points, this analysis was borderline powered to decisively comment on this association. Therefore, no causal analysis was pursued in this effort. Given the observed signals suggesting changes in IOP, along with biological plausibility and alignment with prior preclinical findings, further investigation is warranted to elaborate on the main findings of our study.

Results should be interpreted with the inherent limitations of retrospective studies in mind. We restricted the comparison group to individuals receiving antidiabetic medications to facilitate comparisons of ΔIOP by using a similar index date for both groups. An alternative approach could have been to compare those taking GLP-1R agonists to *all* individuals *not* taking GLP-1R agonists (i.e., include those who are not taking any antidiabetic medications), but this would have presented difficulty in determining corresponding index dates, which could have generated its own bias. Additionally, although no difference was found when comparing the distribution of tonometry recording times between groups, indicating similar measurement times, all IOP records were from daytime clinic visits, and may have missed diurnal variations and peak IOPs. Given these limitations, future prospective studies with standardized protocols are needed to further elucidate the effects of GLP-1R agonists on IOP.

Our study had multiple strengths. This was a retrospective cohort study using the UCHDW, a data repository that includes clinical data of 8.7 million patients across the University of California (UC) health network.^33^ This data warehouse is unique for the purpose of this study in that it is one of the only big data resources that simultaneously includes data on systemic medication usage and IOP values. The Intelligent Research In Sight (IRIS) Registry ^46^ of the American Academy of Ophthalmology includes IOP values but does not have robust data regarding systemic (non-ocular) medications. Other well-known data repositories, such as the National Institutes of Health *All of Us* Research Program,^47^ include systemic medications and clinical information but do not currently have a robust representation of IOP values.^48^ This study is well-powered to evaluate the outcome measure with a sample size of 2330 eyes of 1173 individuals. With an alpha of 0.05 and a beta of 0.8, a sample size of 101 eyes in each group (totaling 202 eyes) would be necessary to detect reductions of ≥1 mmHg. To detect a difference of 0.2 mmHg, the study would require 3771 eyes in each group (totaling 7542 eyes). Given the results of this power analysis, our study falls between these two extremes and is well-powered to state that the difference in effect sizes between the groups is less than 1 mmHg. Another strength of our present study was the statistical analysis, which accounted for potential confounding variables and allowed the generation of robust results. Furthermore, our study was guided by strong preclinical evidence. It stands as one of the initial clinical studies describing the association between GLP-1R agonists exposure and ΔIOP in humans and comparing it to other oral antidiabetics using real-world data. Future prospective studies with study protocol providing for dosage standardization may provide further insights.

## Data Availability

Data availability is controlled by UCOP and not released publicly to preserve privacy of patients. Researchers within the University of California system can contact the relevant party on the link provided below.

https://www.ucop.edu/uc-health/departments/center-for-data-driven-insights-and-innovations-cdi2.html

---

**Supplemental Figure.**
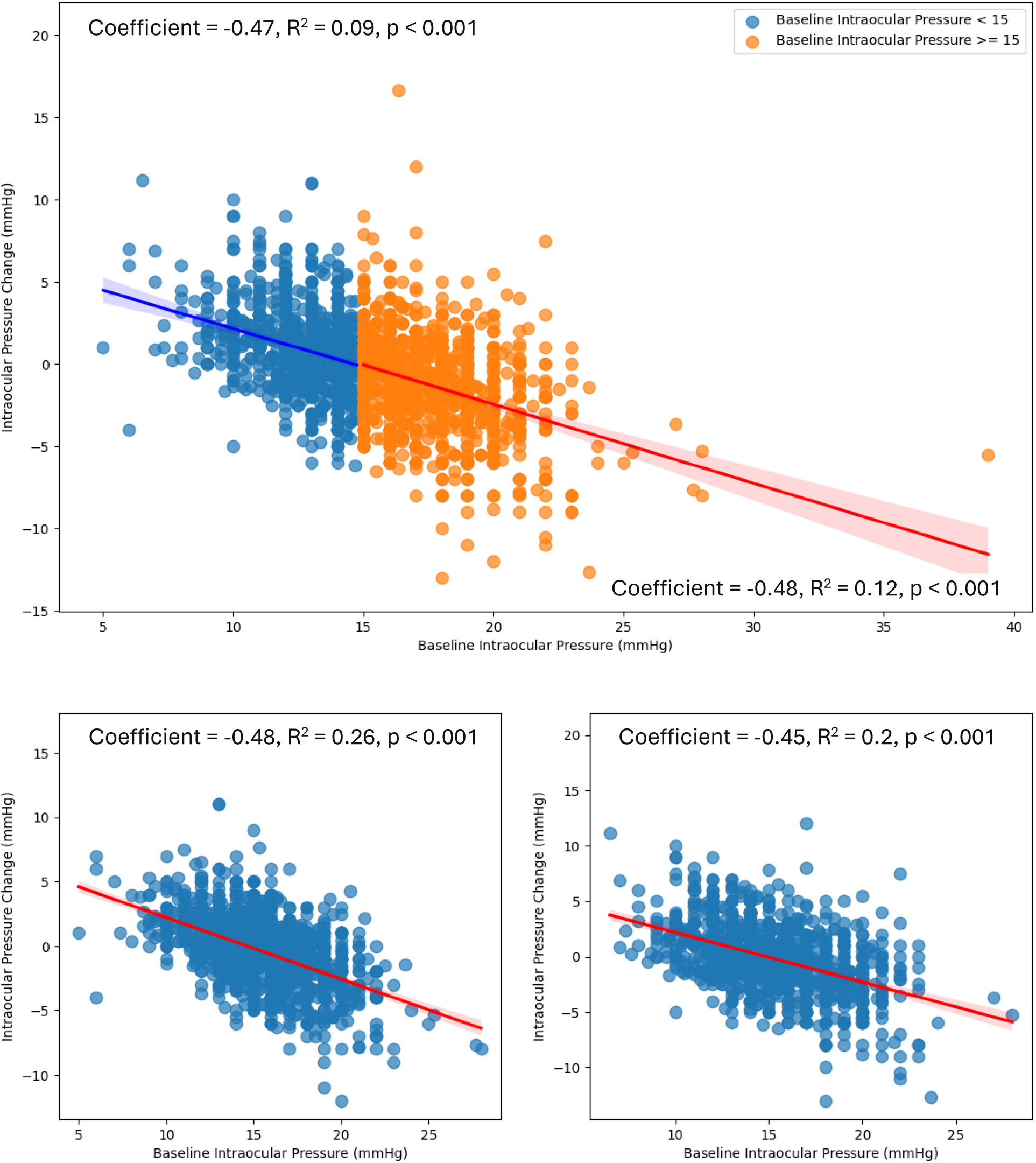
Scatterplot of intraocular pressure changes over pre-exposure intraocular pressures in the entire study population *(top)*, in the glucagon-like peptide 1 receptor agonist group (*bottom left)*, and the control group (*bottom right*).

**Supplementary Table 1:**
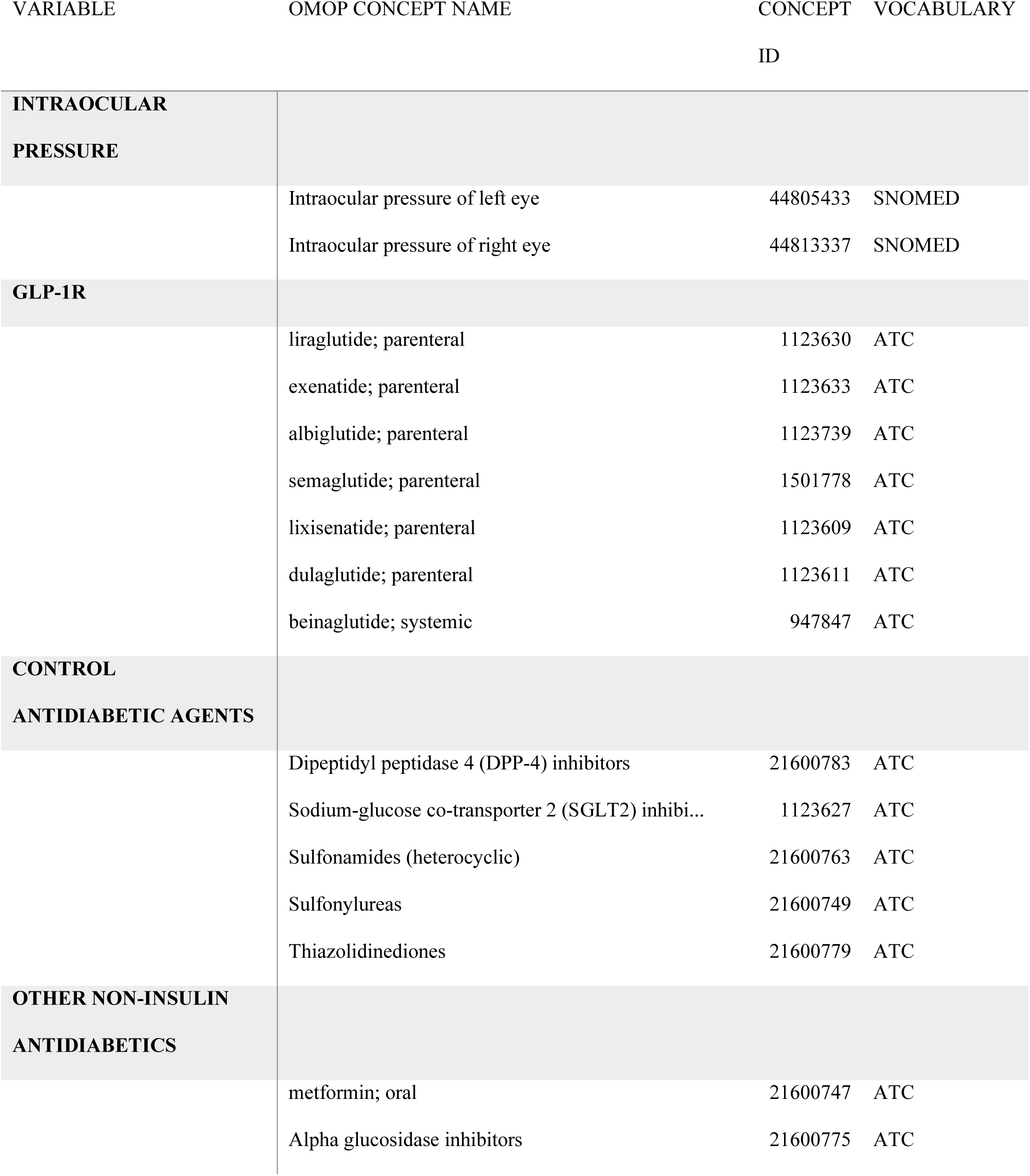
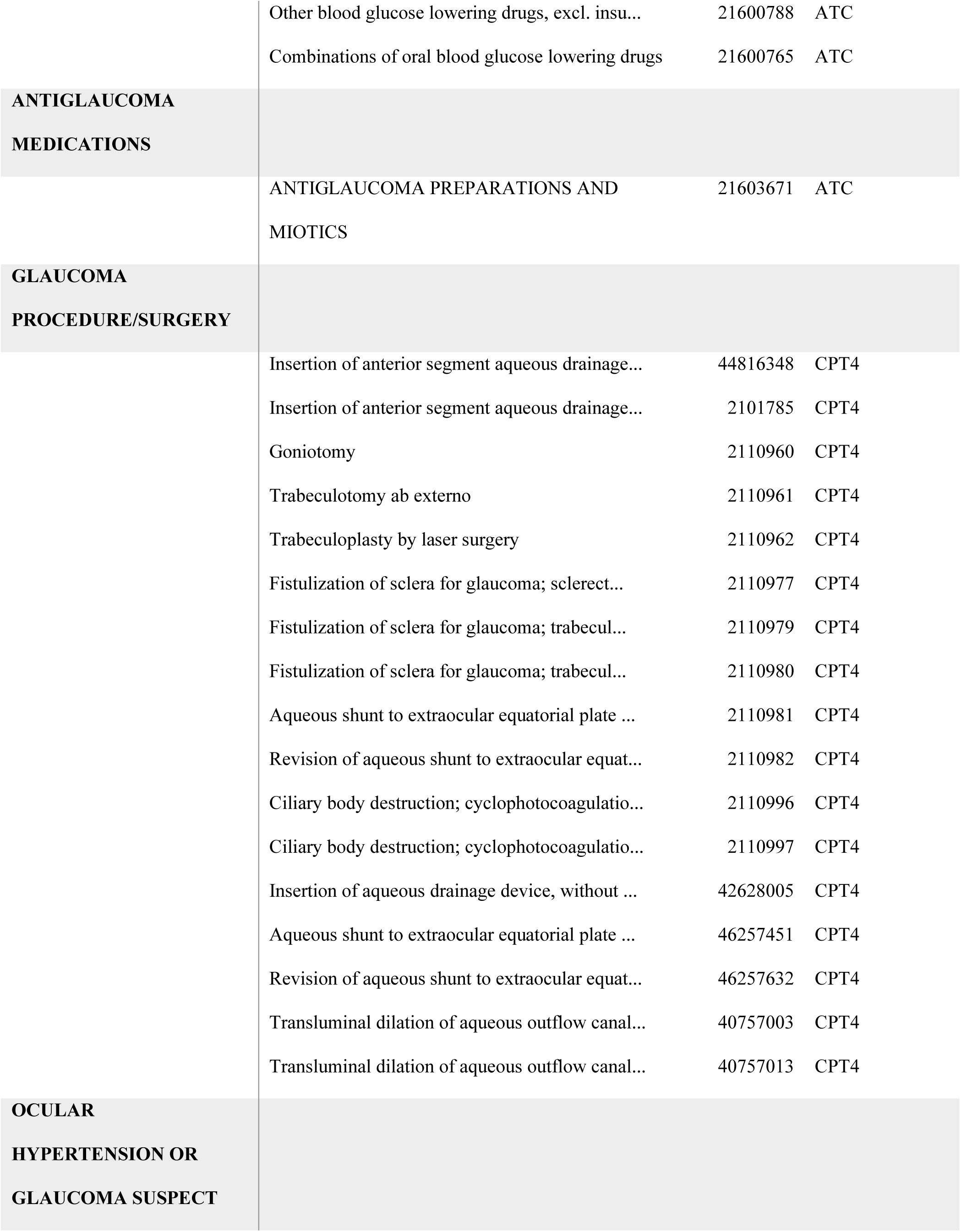
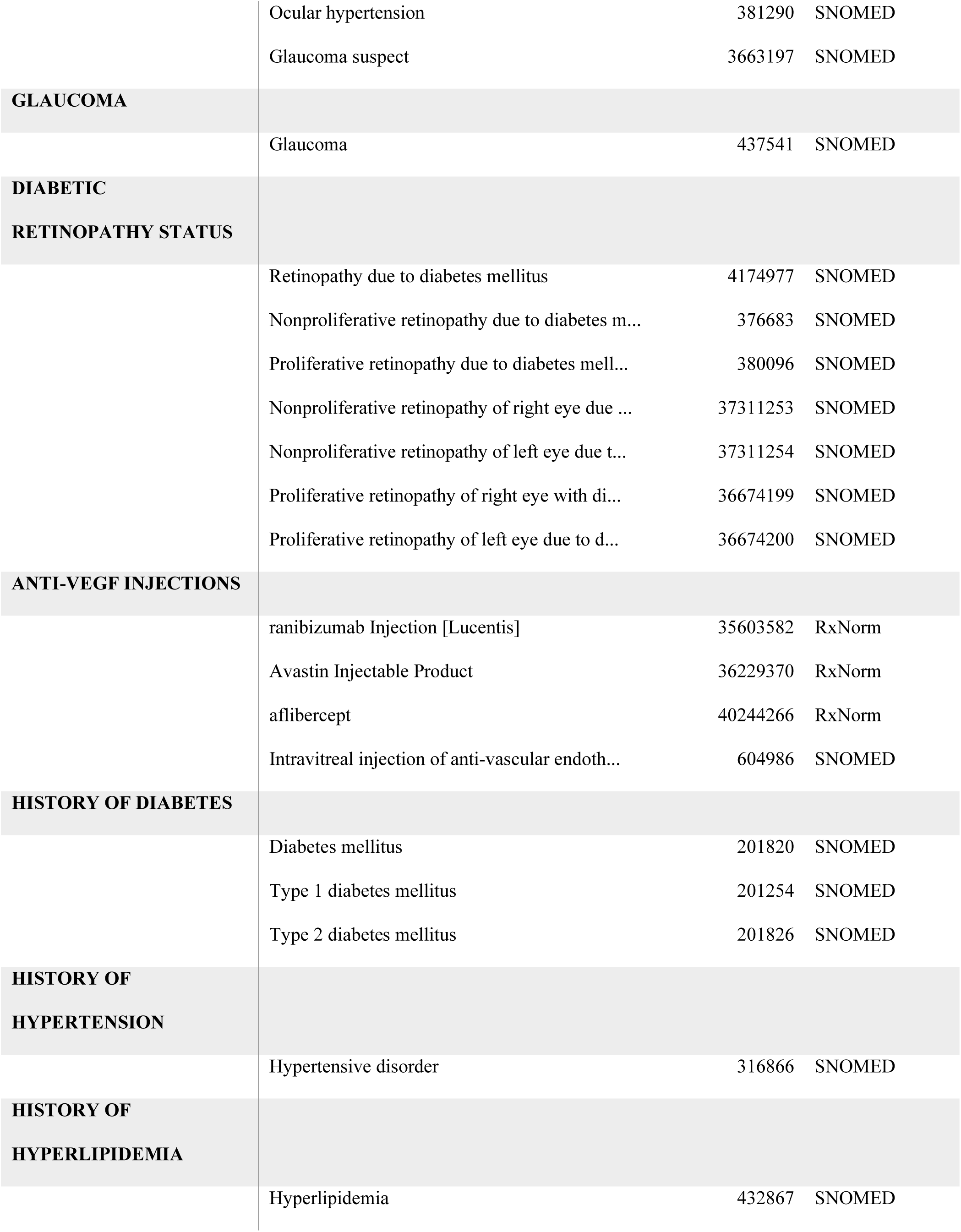
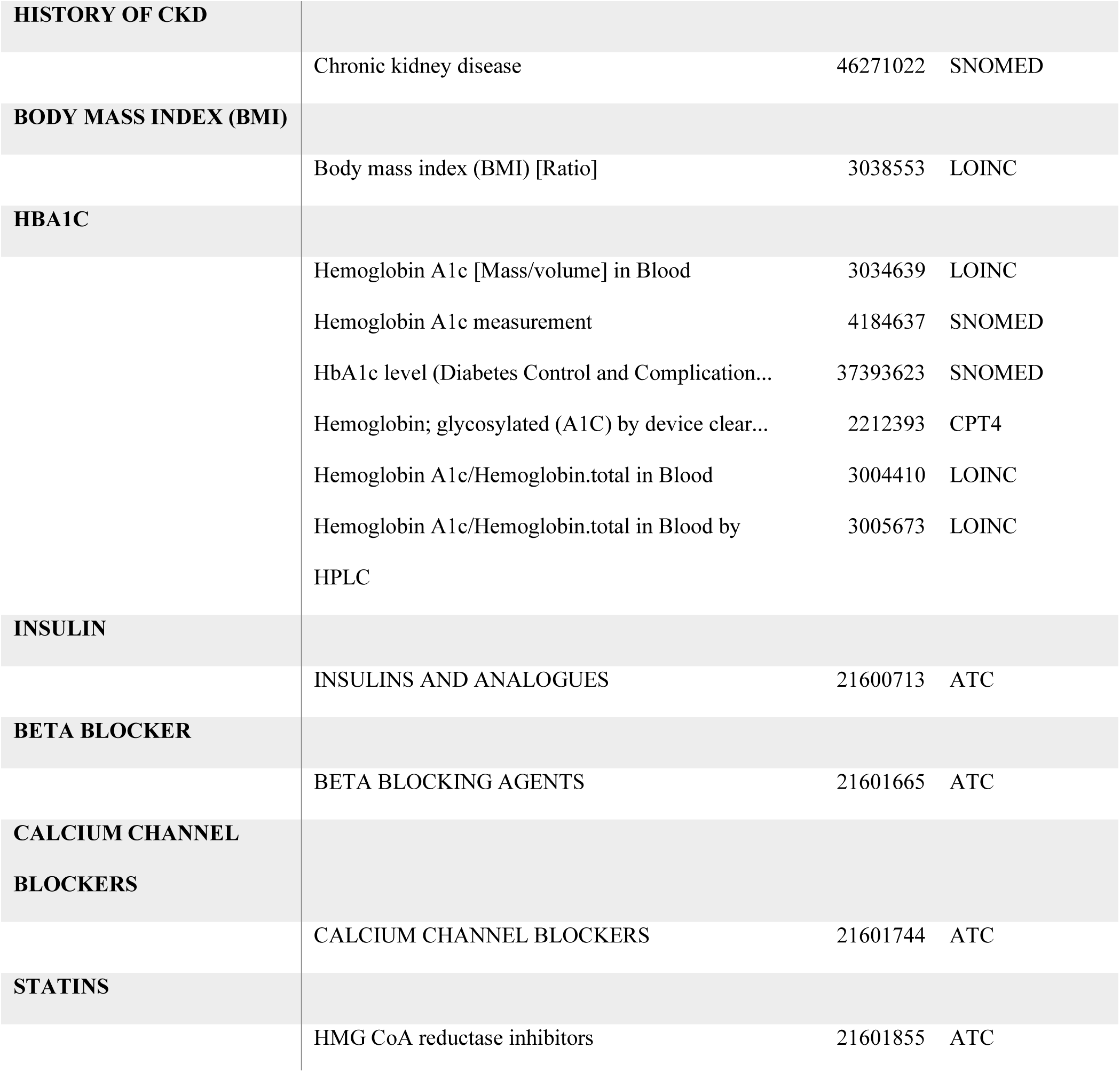
Concept IDs used for identifying records of the variables mentioned in this study. When extracting data, the database was queried for records with concept_ids equal to these codes, or to any descendant in the hierarchy of the OMOP vocabulary. DCSI codes are excluded for clarity, but the codes used are documented in the following repository: https://github.com/OHDSI/FeatureExtraction/blob/bb0213e64198b98e8fe80c3e964d308952779515/inst/sql/sql_server/Dcsi.sql. Acronyms: ATC = Anatomical Therapeutic Chemical; LOINC = Logical Observation Identifiers Names and Codes; CPT4 = Current Procedural Terminology 4; SNOMED = Systematized Nomenclature of Medicine Clinical Terms. RxNorm provides normalized names for clinical drugs.

